# Psychiatric hospital admissions and linkages to ambulatory services in the Western Cape Province of South Africa (2015-2022): trends, risk factors and possible opportunities for intervention

**DOI:** 10.1101/2023.05.17.23290107

**Authors:** Hannah Hussey, Timothy Mountford, Alexa Heekes, Carol Dean, Marinda Roelofse, Lynne Hendricks, Qhama Cossie, Liezel Koen, Warren Cesar, Vanessa Lomas, David Pienaar, Giovanni Perez, Andrew Boulle, Katherine Sorsdahl, Hassan Mahomed

**Author notes:** Contributing author: Hannah Hussey. Author contributions. HH conceptualized the research, performed the formal analysis and visualization, and wrote the original draft. TM & AH were responsible for data curation. CD, LH, MR, GP assisted with the conceptualization of the research. QC, LK, WC, VL, DP and AB assisted with reviewing and editing. KS and HM assisted with reviewing and editing, as well as supervision.

## Abstract

**Background:** Psychiatric hospital admissions in the Western Cape are increasing, driven by poverty and substance use.

**Aim:** To assess the trend of psychiatric admissions from 2015-2022 and factors associated with repeat psychiatric admissions and linkage to ambulatory services post-discharge.

**Setting:** Public hospitals in the Western Cape, South Africa

**Methods:** Using electronic data from the Provincial Health Data Centre, a consolidated routine service database, all psychiatric hospital admissions in the Western Cape were analyzed, stratified by hospital level. Mixed effects logistic regression was used to determine factors associated with successful linkage to ambulatory services within 30 days following hospital discharge, and repeat psychiatric admission within 30 and 90 days.

**Results:** Psychiatric hospital admissions, particularly at the district/acute level, were increasing prior to 2020 and an increasing proportion were substance related. 40% of admissions at the district level had not been seen at a primary health care facility in the year prior to admission. Males and those with substance use disorders were less likely to be successfully linked to outpatient services post-discharge. Successful linkage was most protective against readmission within 90 days with an adjusted odds ratio of 0.76 (95%CI 0.73-0.79) and 0.45 (95%CI 0.42-0.49) at district/acute and specialized hospitals respectively.

**Conclusion:** Improving linkage to ambulatory services for mental health patients post-discharge is likely to avert hospital readmissions.

**Contribution:** This research highlights how often mental health patients requiring admissions are not seen at the primary health care level and quantifies the risk for readmission of not following up psychiatric admissions post-discharge.

## Background

Prior to the COVID-19 pandemic, and consistent with international trends, there was already a high and increasing prevalence of mental health conditions in the Western Cape Province of South Africa (1). Recent surveys in the Western Cape have found that 31.8% of respondents had depression, and that the lifetime prevalence for a substance use disorder was 20.6% (2,3). Nationally, neuropsychiatric disorders are the third highest contributors to disability-adjusted life-years, after HIV/AIDS and other infectious diseases, surpassing other non-communicable diseases (4). Factors such as poverty, food insecurity, experiences of trauma and violence, substance use and comorbid illnesses are all known drivers of this mental health burden (1,3,5–7).

Into this setting, the COVID-19 pandemic occurred, simultaneously exacerbating the burden of mental health disorders, while diverting resources away from mental health care towards COVID-19 services (8– 10). The economic crises and resultant job losses precipitated by COVID-19 also resulted in an increase in depressive symptoms in South Africa (11). High rates of depression, anxiety and post-traumatic stress disorder were also documented in South African healthcare workers during the pandemic (12). In addition, SARS-CoV-2 infection itself can have neuropsychiatric manifestations, including index-episode psychosis (13).

This increase in mental health disorders has particularly impacted in-patient services, where hospital managers have reported struggling to cope with high admission numbers. And while the majority of patients with mental health conditions do not require inpatient care, inpatient expenditure represented 86% of total mental health expenditure in the public health services in South Africa, with specialized psychiatric hospital admissions alone requiring 46% (14). In an increasingly financially constrained environmental, it is crucial to understand how admissions can be reduced and what factors influence this.

This study therefore aimed to describe and interrogate trends in psychiatric hospital admissions from 2015-2022, with a particular focus on the possible impact of substance use and COVID-19. This study will also take into account ambulatory visits before and after admissions, as possible opportunities for interventions.

## Methods

### Setting

The Western Cape Province of South Africa has a population of 7.1 million individuals, 74.9% of whom are reliant on public health services (15). For psychiatric services, the adult population has 33 district hospitals, four regional hospitals and two central hospitals (which we grouped together as “district/acute hospitals” for this analysis), all of whom refer to the three specialised psychiatric hospitals of Lentegeur, Stikland and Valkenberg. In the public sector in South Africa, the inpatient services treatment gap is high at 89%, with only 0.31 psychiatrists and 0.97 psychologists available per 100 000 population (16).

### Data sources

The Provincial Health Data Centre (PHDC) collates all available patient-level electronic health data captured into routine health information systems within public health facilities in the Western Cape province (17). This includes admissions to psychiatric wards, derived from Clinicom, a patient administration database used at all public hospitals in the province, as well as eCCR (electronic Continuity of Care Record), the electronic discharge summary that includes information on discharge diagnoses using ICD-10 coding. Using a unique patient identifier, the admissions are linked to the patients’ interactions with the provincial health system at hospital and primary care levels prior to admission and following discharge.

### Study population and inclusion/exclusion criteria

Individuals of all ages with a psychiatric hospital admission start date from 1 January 2015 to 31 December 2022 in a Western Cape public health facility were included. As ICD-10 diagnostic coding was not always reliable enough to identify psychiatric patients, admitting specialty was used instead. A psychiatric hospital admission was defined as:

- An admission to a specialized psychiatric hospital OR
- An admission to any other hospital in the province, where patients were admitted to the specialty of “Psychiatry” on Clinicom

Patients with psychiatric conditions seen only in the Emergency Centres or those admitted to other specialties (e.g., suicidal patients admitted to the General Medicine Department following an overdose) were not included.

### Data analysis

Firstly, descriptive data looking at trends in admission numbers, diagnoses and outcomes after discharge are presented. First contacts with the public health system, following hospital discharge are also explored. While this is not limited to mental health ambulatory services only, as these can be difficult to distinguish electronically at the primary health care (PHC) level, the assumption is that as all patients are given a follow-up date after discharge, a visit directly after discharge was assumed to be most likely related to the preceding admission. PHC visits in the year preceding a psychiatric admission to an district/acute hospital are also assessed. PHC visits within 24 hours of a hospital admission were excluded here, i.e., those PHC visits where the patient could have been referred on for hospital care were not included, but those PHC visits where the patient went home after and were unrelated to the current hospital admission were. Where appropriate trends are highlighted using Microsoft PowerBI (Version: 2.116).

The second analytical section is restricted to adult general psychiatry patients, as they are the patient population putting the most pressure on services, discharged home from either a district/acute hospital or a specialized hospital, i.e., admissions that result in transfers from a district/acute hospital to a specialized hospital are excluded. Elective therapeutic admissions to specialized hospitals were also excluded. Any individuals who were known to be deceased (in-facility deaths of individuals with a previous psychiatric admission) were also excluded. Stata (13.1, StataCorp. LLC, College Station, TX) was used for the analysis. The outcomes assessed were:

- Successful linkage to care post-discharge, defined as an ambulatory visit (PHC clinic or hospital out-patient) within 30 days of hospital discharge.
- Repeat psychiatric hospital admission, at any hospital level, within 30 and 90 days from the date of discharge

All admissions discharged by 31 December 2022 were included in this analysis (the data set was pulled on 15 March 2023, allowing sufficient time for the outcomes to have occurred and been captured electronically), except for the outcome of readmissions within 90 days, where only admissions discharged by 30 November 2022 be included.

Mixed effects logistic regression is used, as individuals often have repeated admissions over time and there is non-independence of observations. An alternative approach of using standard logistic regression, but including only the latest admission for an individual, was also performed. All analyses are stratified by hospital level (district/acute and specialized) and adjusted for:

- Age and sex
- Broad diagnosis category - based on primary discharge diagnosis ICD-10 coding: mood disorders, including both uni- and bipolar depression (F30-F39); schizophrenia and other psychotic disorders (F20-F29); substance related disorders (F10-F19); and other diagnoses

`O` As the coding at specialized hospitals is more detailed, having a substance related diagnosis listed anywhere on the discharge summary (i.e., not just the primary diagnosis) was also included
- Index psychiatric admission or a known previous psychiatric admission, according to the available Clinicom data. Hospitals started using Clinicom at different times from 2001, with the psychiatric hospitals starting in 2008. Index admission data may not be available electronically for patients with longstanding psychiatric disorders
- PHC visit in year preceding admission or not
- Length of hospital stay,
- Admission year
- Admitting hospital

`O` The three specialized hospitals, labelled as A, B and C
`O` The district/acute hospitals grouped into rural regional or district hospitals, and the metropolitan hospitals grouped according to their specialized hospital catchment area (A, B or C)

## Ethics

This study was approved by the University of Cape Town Health Research Ethics Committee (HREC 058 /2023). Informed consent was waived for this secondary analysis of de-identified data.

## Results

### General trends in increasing admission numbers

Psychiatric admissions in the province have increased markedly over the last eight years (Figure 1a). In 2015 there were 6 013 admissions in specialized hospitals and 10 991 admissions in district/acute hospitals. This increased to 6 697 and 19 880 admissions respectively in 2022. Admissions in district/acute level hospitals almost doubled during this period, while specialized hospitals only increased slightly.

**Figure 1:**
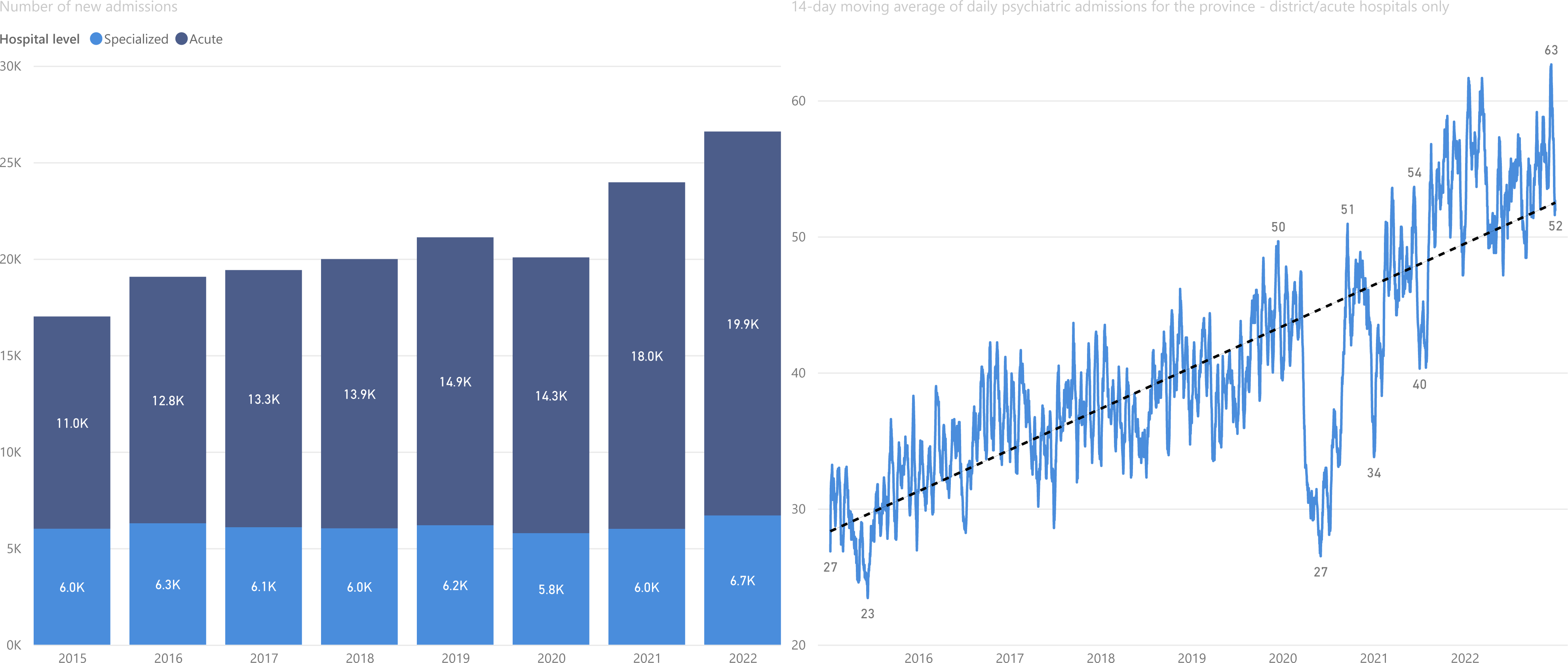
**Psychiatric hospital admissions in the Western Cape 2015-2022 - annual admissions for all facilities (1a) and 14 day moving average of psychiatric admissions at acute/district hospital level 2015-2022 (1b).**

2020 was the only year where an increase in admissions was not seen. For district/acute admissions (Figures 1b), we can see a large decrease in admission numbers coinciding with the first wave of COVID-19 infections in the province in mid-2020, followed by smaller decreases with the second (December 2020 – January 2021) and third waves (mid-2021).

Figure 2b illustrates the primary discharge diagnosis in specialized hospitals for general acute psychiatry, where schizophrenia and other psychotic disorders make up more than half the primary diagnoses, with mood disorders contributing to less than 20% of admissions. Substance related disorders as the primary discharge diagnosis has increased as a proportion from 15.96% in 2018 to 22.47% in 2022. When looking at all diagnosis codes listed (i.e., not just the primary diagnosis code) (Figure 2d), substance related diagnoses also increased proportionately with time, accounting for 45.84% of all admissions in 2022.

**Figure 2:**
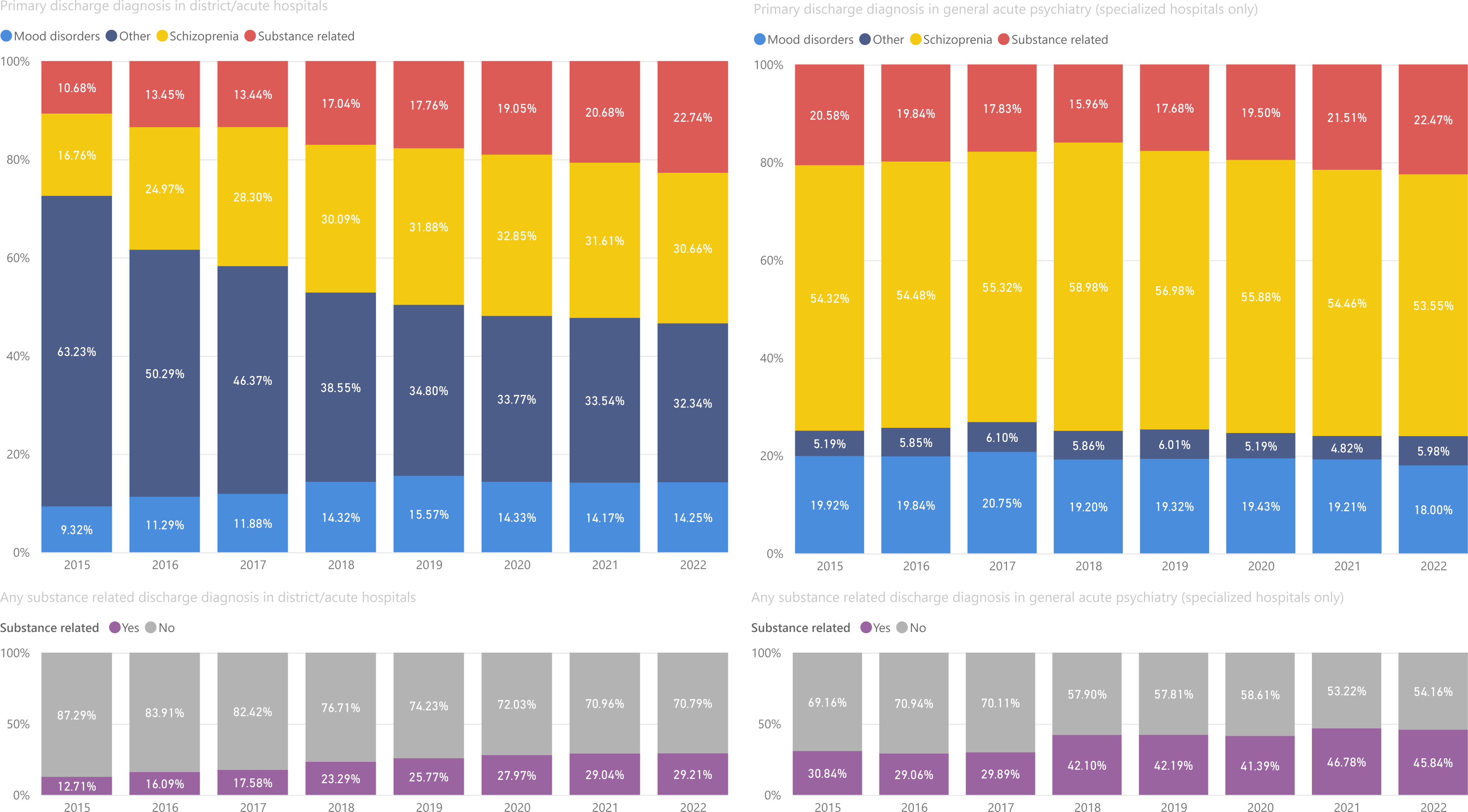
**Discharge diagnosis trends for acute/district hospitals (2a) and specialized hospitals (2b), based on the primary discharge diagnosis. The proportion of admissions that had any substance related ICD-10 code listed at acute/district hospitals (2c) and specialized hospitals (2d).**

Diagnostic coding is not as detailed or complete at district/acute hospitals, although this has improved with time – as illustrated by the initially large number of diagnoses that fell into the “Other” category. But a similar trend can be seen at these hospitals (Figures 2a and 2c), where substance related disorders now make up 22.74% of all primary diagnoses in 2022, and 29.21% of all admissions have a substance related diagnosis listed in 2022. While the proportion that were substance related increased over time, so did the total absolute number of patients too, meaning that the 12.71% of patients with any substance related diagnosis code in 2015 corresponded to 1 396 individuals and the 29.21% of patients in 2022 corresponded to 5 613 individuals.

The readmission rate has, however, remained relatively stable, with around 10% of admissions, resulting in a psychiatric readmission in the next 30 days (Figure 3), and this increases to around 18-19% at 90 days. But with the absolute number of psychiatric hospital admissions increasing, so too have the absolute number of readmissions putting the limited beds available under pressure. Currently only 47.5% of admissions acute/district hospitals and 28.68% of admissions to specialised hospitals are first known psychiatric admissions. Amongst those known with to have a previous psychiatric admission, the median number of days between admissions decreased between 2018 to 2022, for both hospital levels.

**Figure 3:**
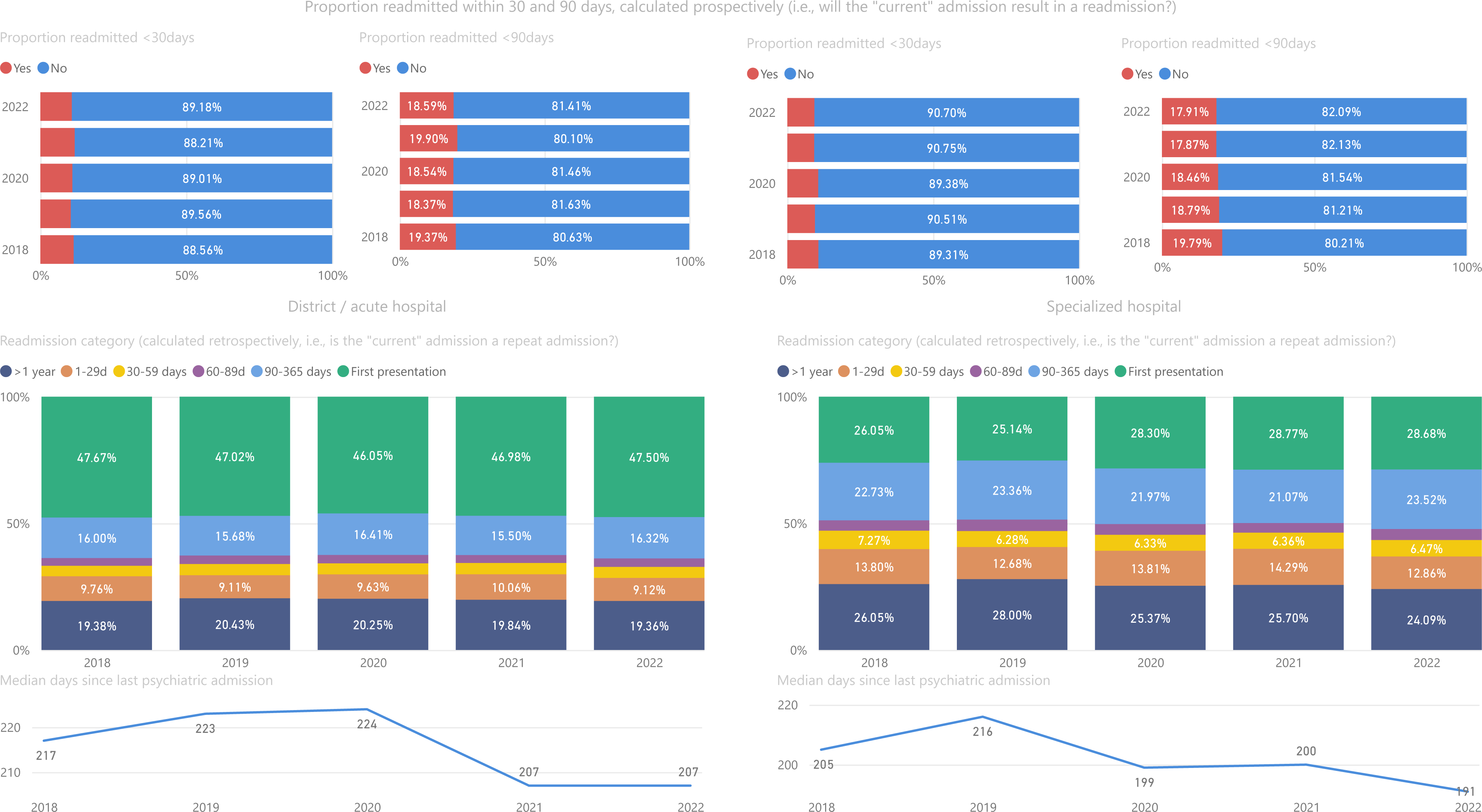
**Readmission rates for district/acute and specialized hospitals 2018-2022 - readmission within 30 days (3a), 90 days (3b), readmission categories (3c) and median days between psychiatric admissions (3d).**

### Profile of admitted patients and outcomes after discharge

Male patients account for 62.15% of all psychiatric admissions in the province. Male patients and those with a primary substance related disorder tended to be younger (Supplementary Figure 1).

In 2022, the median length of stay in district/acute hospitals was 7 days and for specialized hospitals 34 days. In district/acute hospitals, those awaiting referral required an additional week of admission (Supplementary Figure 2a). With increasing numbers of admissions at the district/acute level, a decreasing proportion were referred on for specialist care. In 2015, 37.54% were referred on, while in 2022 only 27.08% were referred on. Length of stay at specialised hospitals was related to discharge diagnosis (Supplementary Figure 2b). Schizophrenia and other psychotic disorders had the highest lengths of stay, followed by mood disorders and then substance related disorders. All diagnosis categories showed a decrease in length of stay for 2022.

A median of 12 and 16 days were taken to first contact with the health system after discharge from district/acute hospitals and specialized hospitals, respectively. For those discharged directly from a district/acute hospital in 2022, 53.2% of individuals had an clinic/out-patient visit within 30 days, a number which has been increasing slowly over the last few years (Figure 4a). An additional 10.43% do make contact with out-patient services but take >30 days after discharge. 8.37% of discharges only make contact with the health system again with their next admission and 28.0% have had no further contact with the health system (as of March 2023, when this dataset was extracted). A similar trend is seen at specialized hospitals, although a higher proportion are followed up at ambulatory services within 30 days – 65.4% in 2022 (Figure 4b). But around 20% of patients discharged from specialized psychiatric service have no further contact with the health system.

**Figure 4:**
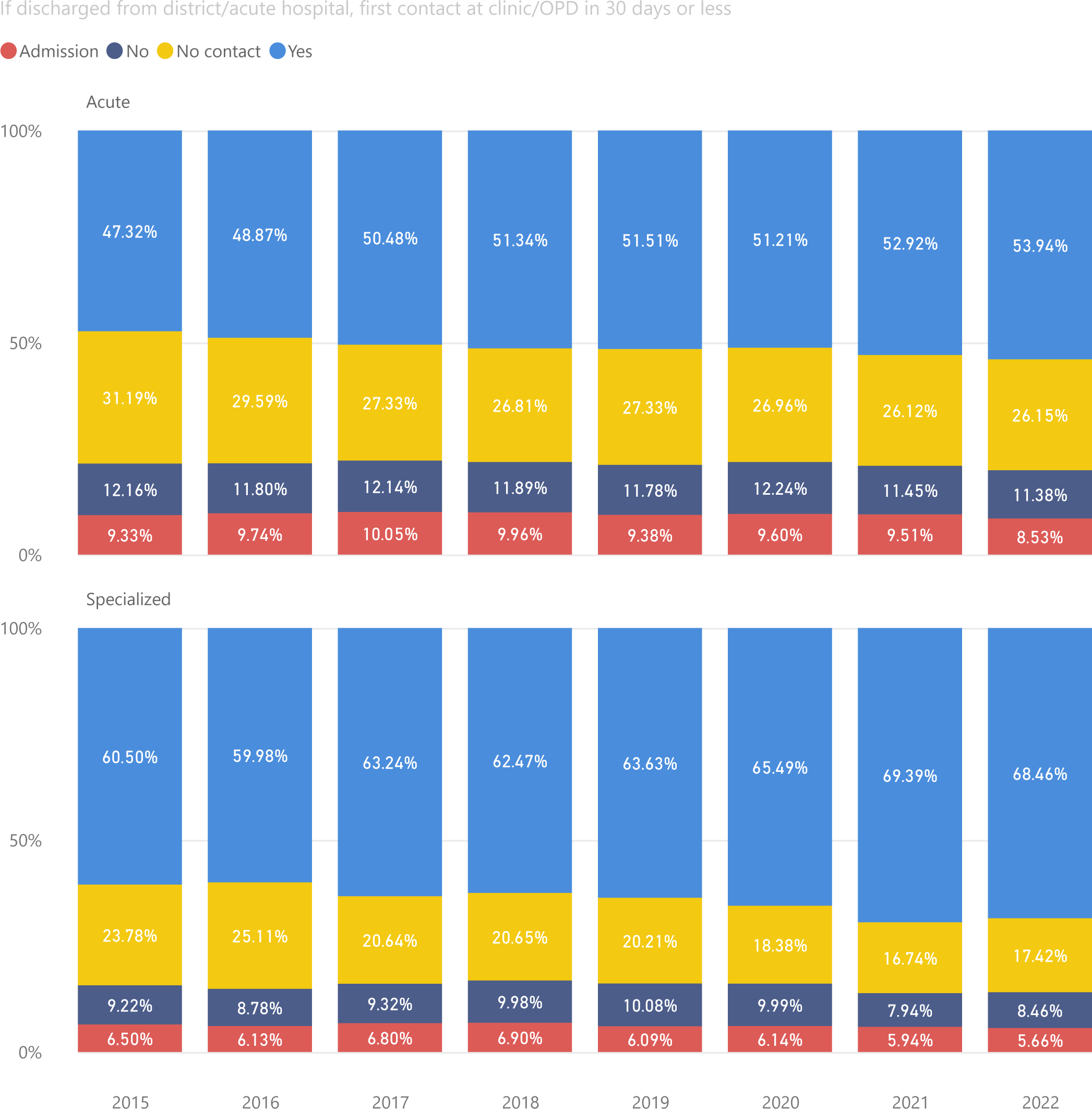
**After discharge from hospital, the clinic or out-patient contact within 30 days is assessed at acute/district hospitals (4a) and specialized hospitals (4b). “Yes” is defined as an ambulatory (clinic or out-patient) visit within 30 days, while “No” is an ambulatory visit but at more than 30 days.**

### PHC visits before a psychiatric admission

In 2022, 39.4% of admitted patients had not been seen at a PHC facility in the year preceding their admission, and this increased to 72.35% if one looked at the 30 days preceding an admission (Supplementary Figure 3). Since 2015 there has been a general trend of increasing PHC visits in the period before an admission. 2020, however, was the exception with the proportion of admissions with a PHC visit in the 30 days prior to admission decreasing in May to July. Despite this general increase, the median days between the last PHC visit and the psychiatric admission has remained relatively stable between 35 and 42 days (37 days in 2022).

### Successful linkage to care

74 728 admissions at district/acute hospitals and 30 750 admissions at specialized hospitals were included in these analyses, the detailed characteristics of which are described in Table 1.

**Table 1:**
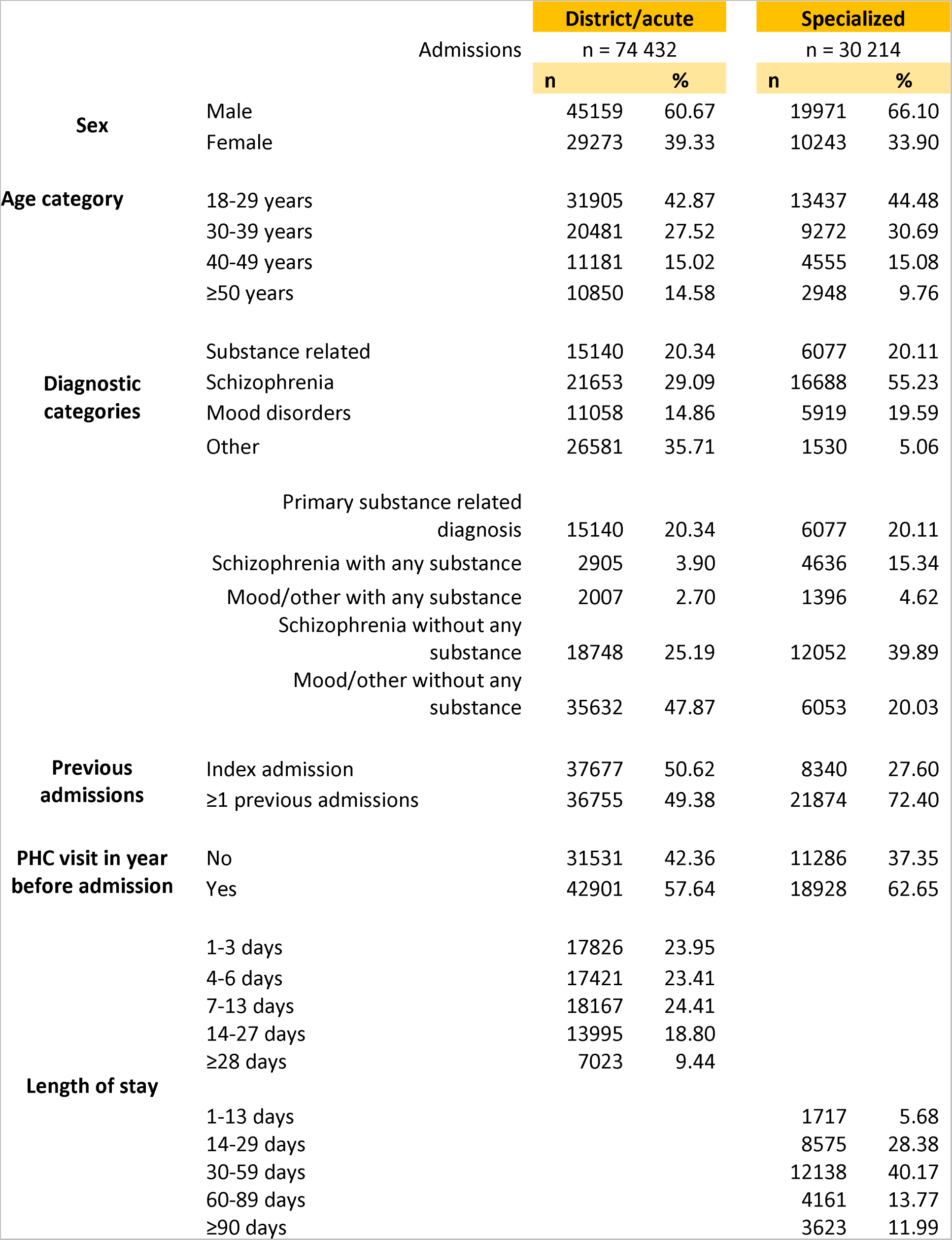
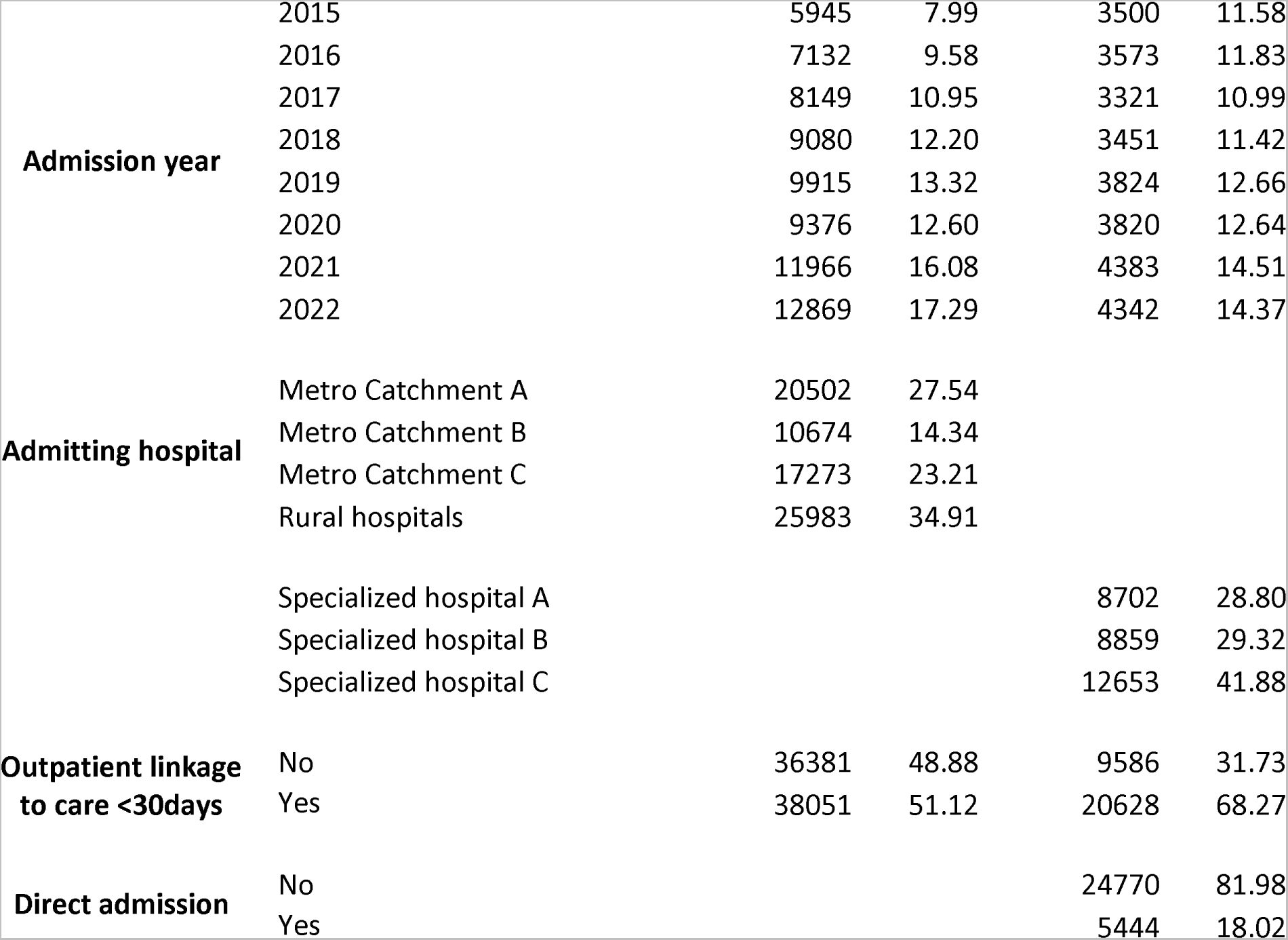
Detailed characteristics of included admissions, stratified according to hospital level.

At district/acute hospitals (Table 2), those with a PHC visit in the preceding year were more likely to be successfully linked to ambulatory services (adjusted Odds ratio [aOR] of 2.67 [95% confidence interval {95%CI} 2.58-2.77]). Women were also more likely to be successfully linked to care, as were older patients and those without a primary substance related disorder. At specialized hospitals (Table 3) similar trends were seen, bearing in mind the slightly different categorization of diagnoses and lengths of stay. Those with a PHC visit in the year prior to admission (aOR 2.07 [95%CI 1.93-2.22]) and women (aOR 1.14 [95%CI 1.05-1.23]) were more likely to be successfully linked to care, but increasing age and previous psychiatric admissions were not associated with improved linkage to care.

**Table 2:**
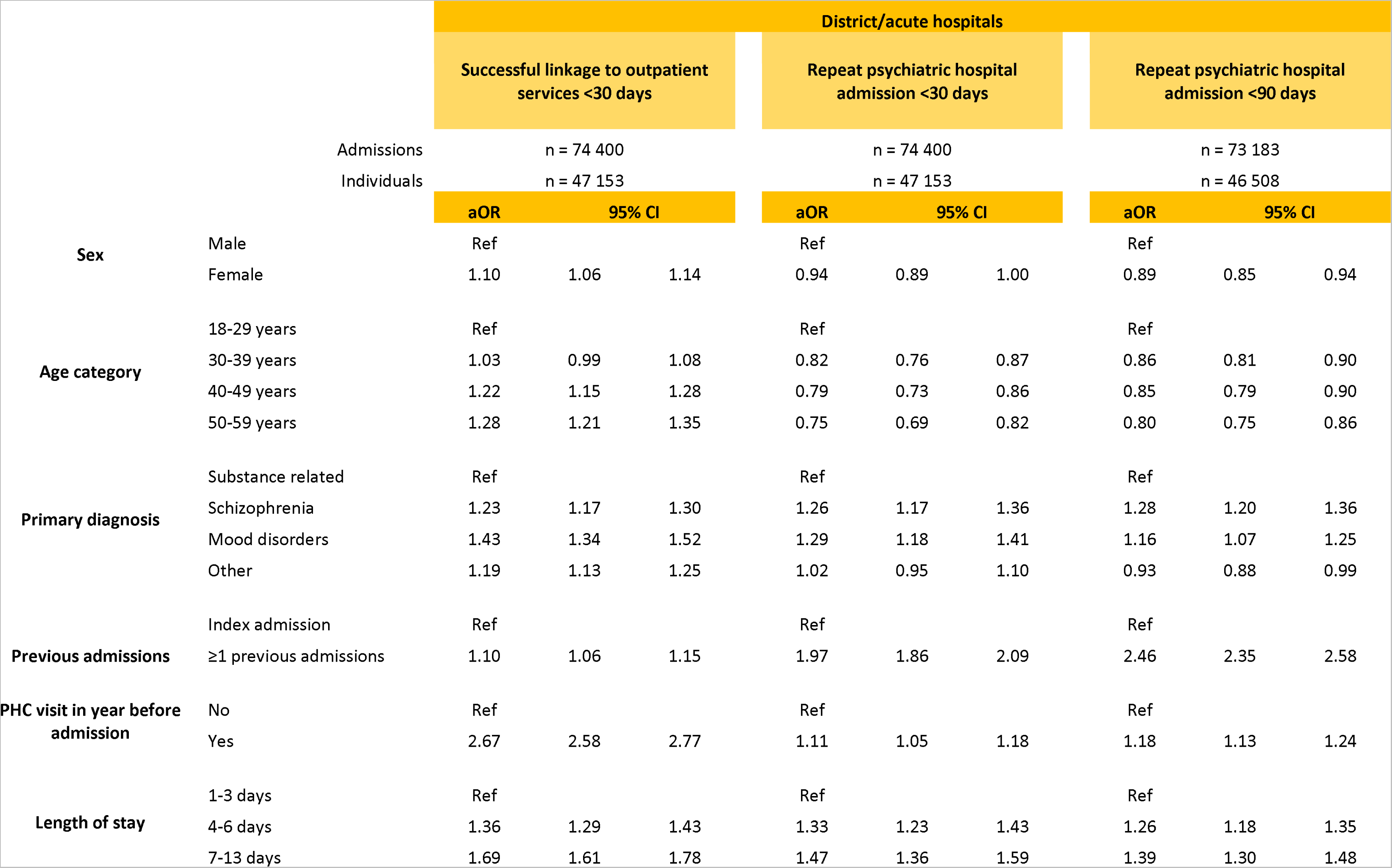
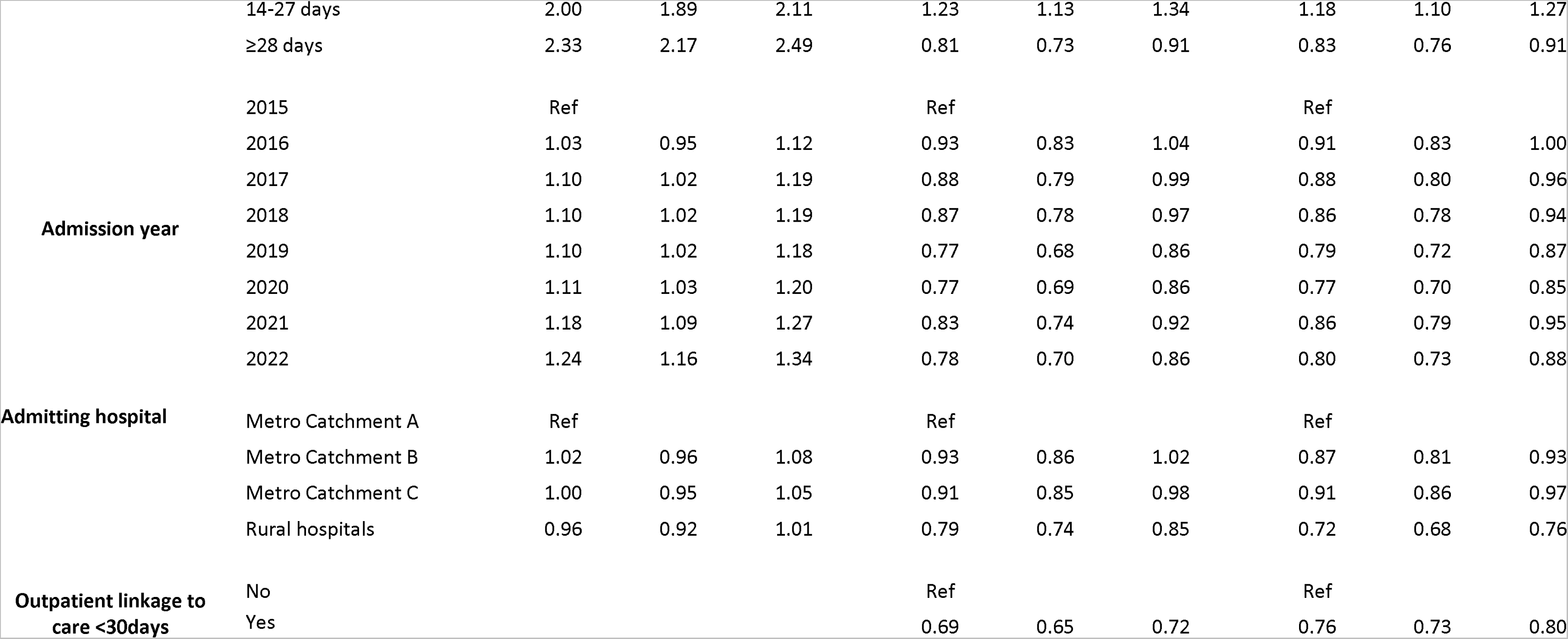
Mixed effects logistic regression for the outcome of successful linkage to care following discharge (i.e., a clinic or out-patient visit within 30 days of hospital discharge) and psychiatric readmissions within 30 and 90 days for district/acute hospitals. Only patients discharged directly home are included in these analyses.

**Table 3:**
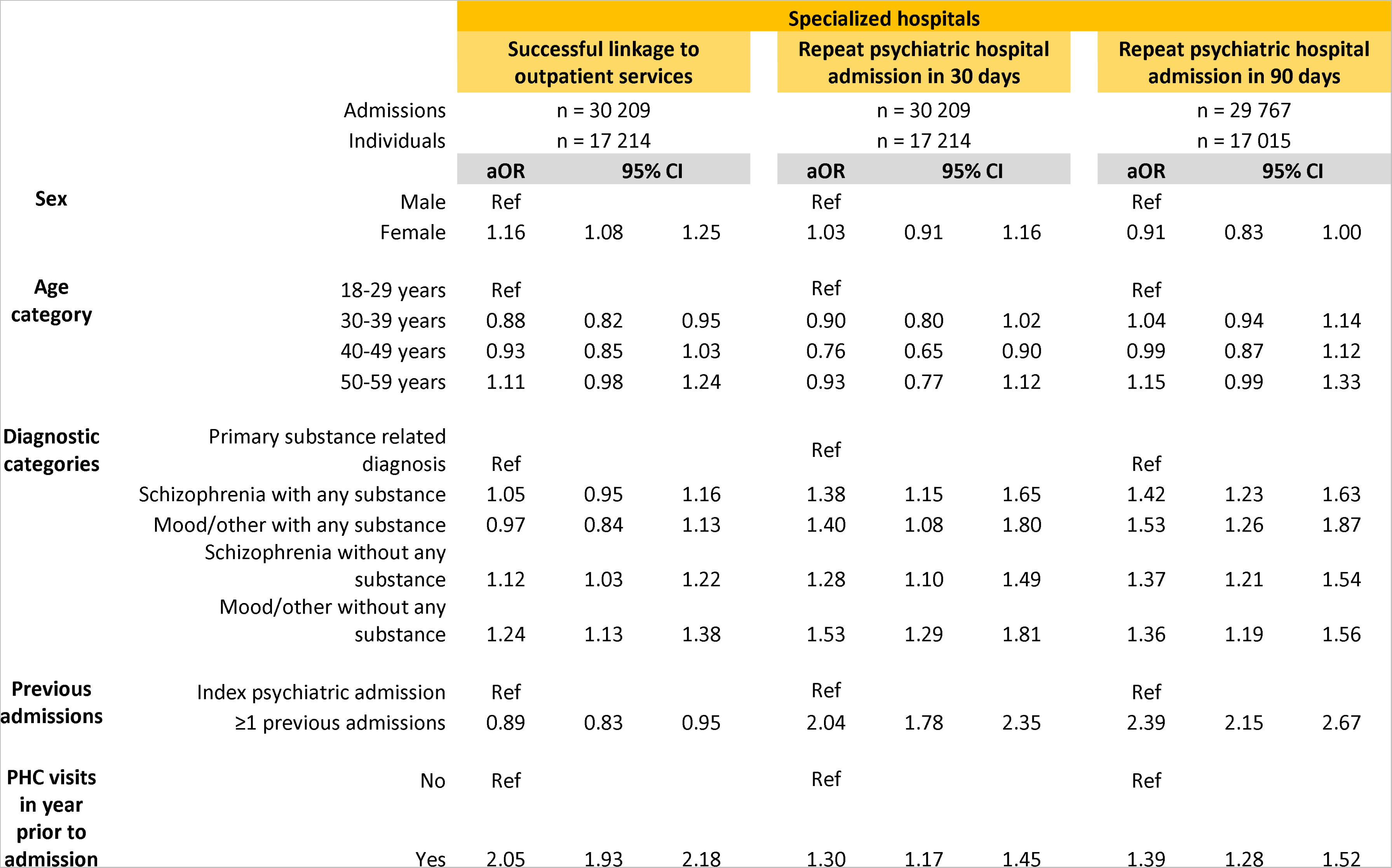
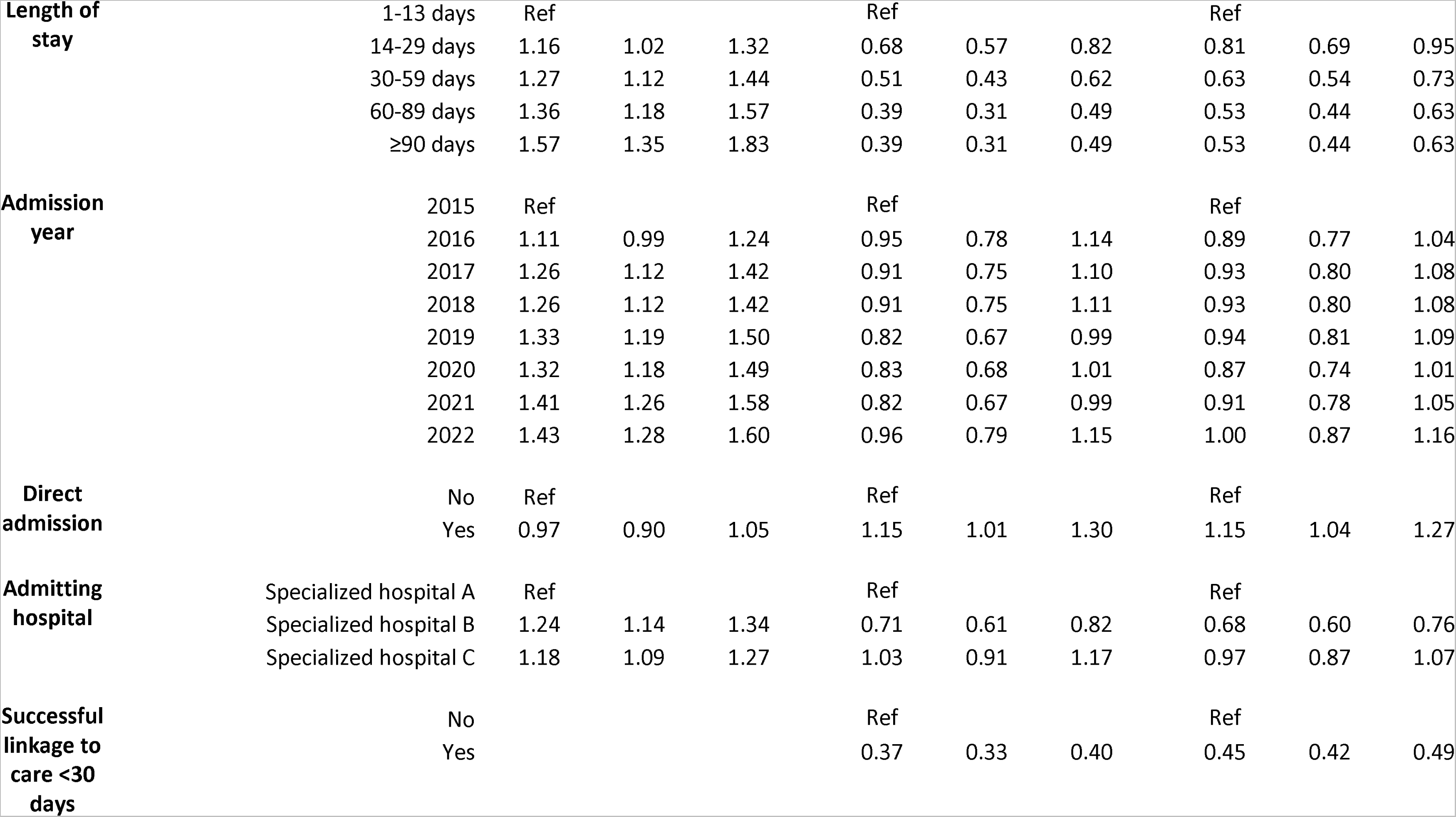
Mixed effects logistic regression for the outcome of successful linkage to care following discharge (i.e., a clinic or out-patient visit within 30 days of hospital discharge) and psychiatric readmissions within 30 and 90 days for specialized hospitals.

|  |  | Specialized hospitals |  |  |  |  |  |  |  |  |
| --- | --- | --- | --- | --- | --- | --- | --- | --- | --- | --- |
|  |  | Successful linkage to outpatient services |  |  | Repeat psychiatric hospital admission in 30 days |  |  | Repeat psychiatric hospital admission in 90 days |  |  |
|  |  | Admissions<br>n = 30 209 |  |  | n = 30 209 |  |  | n = 29 767 |  |  |
|  |  | Individuals<br>n = 17 214 |  |  | n = 17 214 |  |  | n = 17 015 |  |  |
|  |  | aOR | 95% CI |  | aOR | 95% CI |  | aOR | 95% CI |  |
| Sex | Male | Ref |  |  | Ref |  |  | Ref |  |  |
|  | Female | 1.16 | 1.08 | 1.25 | 1.03 | 0.91 | 1.16 | 0.91 | 0.83 | 1.00 |
| Age category | 18-29 years | Ref |  |  | Ref |  |  | Ref |  |  |
|  | 30-39 years | 0.88 | 0.82 | 0.95 | 0.90 | 0.80 | 1.02 | 1.04 | 0.94 | 1.14 |
|  | 40-49 years | 0.93 | 0.85 | 1.03 | 0.76 | 0.65 | 0.90 | 0.99 | 0.87 | 1.12 |
|  | 50-59 years | 1.11 | 0.98 | 1.24 | 0.93 | 0.77 | 1.12 | 1.15 | 0.99 | 1.33 |
| Diagnostic categories | Primary substance related diagnosis | Ref |  |  | Ref |  |  | Ref |  |  |
|  | Schizophrenia with any substance | 1.05 | 0.95 | 1.16 | 1.38 | 1.15 | 1.65 | 1.42 | 1.23 | 1.63 |
|  | Mood/other with any substance | 0.97 | 0.84 | 1.13 | 1.40 | 1.08 | 1.80 | 1.53 | 1.26 | 1.87 |
|  | Schizophrenia without any substance | 1.12 | 1.03 | 1.22 | 1.28 | 1.10 | 1.49 | 1.37 | 1.21 | 1.54 |
|  | Mood/other without any substance | 1.24 | 1.13 | 1.38 | 1.53 | 1.29 | 1.81 | 1.36 | 1.19 | 1.56 |
| Previous admissions | Index psychiatric admission | Ref |  |  | Ref |  |  | Ref |  |  |
|  | ≥1 previous admissions | 0.89 | 0.83 | 0.95 | 2.04 | 1.78 | 2.35 | 2.39 | 2.15 | 2.67 |
| PHC visits in year prior to admission | No | Ref |  |  | Ref |  |  | Ref |  |  |
|  | Yes | 2.05 | 1.93 | 2.18 | 1.30 | 1.17 | 1.45 | 1.39 | 1.28 | 1.52 |

|  |  |  |  |  |  |  |  |  |  |  |  |
| --- | --- | --- | --- | --- | --- | --- | --- | --- | --- | --- | --- |
|  | **Length of stay** | 1-13 days | Ref |  |  | Ref |  |  | Ref |  |  |
|  |  | 14-29 days | 1.16 | 1.02 | 1.32 | 0.68 | 0.57 | 0.82 | 0.81 | 0.69 | 0.95 |
|  |  | 30-59 days | 1.27 | 1.12 | 1.44 | 0.51 | 0.43 | 0.62 | 0.63 | 0.54 | 0.73 |
|  |  | 60-89 days | 1.36 | 1.18 | 1.57 | 0.39 | 0.31 | 0.49 | 0.53 | 0.44 | 0.63 |
|  |  | ≥90 days | 1.57 | 1.35 | 1.83 | 0.39 | 0.31 | 0.49 | 0.53 | 0.44 | 0.63 |
|  | **Admission year** | 2015 | Ref |  |  | Ref |  |  | Ref |  |  |
|  |  | 2016 | 1.11 | 0.99 | 1.24 | 0.95 | 0.78 | 1.14 | 0.89 | 0.77 | 1.04 |
|  |  | 2017 | 1.26 | 1.12 | 1.42 | 0.91 | 0.75 | 1.10 | 0.93 | 0.80 | 1.08 |
|  |  | 2018 | 1.26 | 1.12 | 1.42 | 0.91 | 0.75 | 1.11 | 0.93 | 0.80 | 1.08 |
|  |  | 2019 | 1.33 | 1.19 | 1.50 | 0.82 | 0.67 | 0.99 | 0.94 | 0.81 | 1.09 |
|  |  | 2020 | 1.32 | 1.18 | 1.49 | 0.83 | 0.68 | 1.01 | 0.87 | 0.74 | 1.01 |
|  |  | 2021 | 1.41 | 1.26 | 1.58 | 0.82 | 0.67 | 0.99 | 0.91 | 0.78 | 1.05 |
|  |  | 2022 | 1.43 | 1.28 | 1.60 | 0.96 | 0.79 | 1.15 | 1.00 | 0.87 | 1.16 |
|  | **Direct admission** | No | Ref |  |  | Ref |  |  | Ref |  |  |
|  |  | Yes | 0.97 | 0.90 | 1.05 | 1.15 | 1.01 | 1.30 | 1.15 | 1.04 | 1.27 |
|  | **Admitting hospital** | Specialized hospital A | Ref |  |  | Ref |  |  | Ref |  |  |
|  |  | Specialized hospital B | 1.24 | 1.14 | 1.34 | 0.71 | 0.61 | 0.82 | 0.68 | 0.60 | 0.76 |
|  |  | Specialized hospital C | 1.18 | 1.09 | 1.27 | 1.03 | 0.91 | 1.17 | 0.97 | 0.87 | 1.07 |
|  | **Successful linkage to care <30 days** | No |  |  |  | Ref |  |  | Ref |  |  |
|  |  | Yes |  |  |  | 0.37 | 0.33 | 0.40 | 0.45 | 0.42 | 0.49 |

### Risk factors for readmission

At district/acute hospitals, female sex, older ages, being admitted to a rural hospital and a primary substance related diagnosis were associated with a lower risk of readmission. An increasing length of stay (but only up until 28 days), previous psychiatric admissions and a PHC visit in the year preceding an admission were risk factors for readmission. While the aORs for admission years decreased with time, a relative increase was seen in 2021, although 2022 has seen a return to aORs of pre-pandemic years. Successful linkage to care decreased the risk of readmission by 31% (aOR = 0.69 [95%CI 0.65-0.72]) and 24% (aOR = 0.76 [0.73-0.79]) for readmissions within 30 and 90 days respectively.

At specialized hospitals, successful linkage to care within 30 days of discharge was the most protective factor against readmission within 30 and 90 days, with an aOR of 0.37 (95%CI 0.32-0.43) and 0.49 (0.44-0.55) respectively. Sex was not associated with readmission but a primary diagnosis that was substance related had the lowest association with readmission. Previous admissions were strongly associated with readmissions, as well as a PHC visit in the preceding year and being a direct admission to a specialized hospital (i.e., patients not transferred from a district/acute hospital), although the latter two factors only to a lesser degree. Longer lengths of stay were also protective of readmission, although the aOR for 60-90 days and ≥ 90 days similar. There was no clear trend for admission years.

The alternative approach of using standard logistic regression on only the latest hospital admission for an individual found similar results (Supplementary Table 1 and 2).

## Discussion

The main finding of this study is that a high proportion individuals who require psychiatric hospital admission, are not routinely attending at ambulatory services, both before and after the admission, suggesting that opportunities to avert psychiatric hospital admissions and readmissions are being missed. Substance related disorders are contributing to not only the increase in admission numbers being seen, but once an individual is discharged home, those with a substance related disorder are less likely to successfully link to ambulatory services.

While COVID-19 and its after effects are likely contributing to the current increase in psychiatric admissions, it must be highlighted that the trend of increasing admissions, particularly at the district/acute level, was already established prior to 2020, implying that other factors, such as increasing poverty, inequality, substance use and trauma/violence, could be driving this increase (18). Food insecurity and neighborhood level deprivation were found to be associated with depression, suggesting that structural poverty was contributing to the burden of mental illness in the South Africa (5,6). A study conducted in Lentegeur Hospital in 2016 found that 62% of patients, and 73% of male patients, used substances prior to their admission, with cannabis and methamphetamine being most commonly used (7). While the substance rates we found were lower, they were dependent on clinician ICD-10 coding, which is often not detailed enough, especially at the district/acute hospital level.

Increased admission numbers can also be related to increased access to in-patient health services, particularly in settings where out-patient services are limited. The district/acute hospitals with the highest number of psychiatric admissions in 2022 were the new large district hospitals of Mitchells Plain and Khayelitsha, which opened in 2013 and 2012 respectively. And the increasing size of the general population over time should also not be forgotten (18). Admission numbers at specialized hospitals, however, showed only a muted increase in numbers, in line with provincial policy which preferentially funded the expansion of beds at the district/acute level (Circular H18/2015). According to a Provincial Database, the number of official psychiatry beds in the Cape Town Metro was 417 in 2022 (a 136% increase from 2015), while the three specialized hospitals only increased by 11% in the same time period, to 1 527 beds in 2022.

24% of mental health inpatients nationally are being readmitted to hospital within three months of being discharged (14). Our study found slightly lower rates of 18-19% within 3 months, and no increase in the readmission rate, except for the relative increase in district/acute hospitals in 2021, although with rising absolute numbers of admissions this might still change. Scarcity of in-patient psychiatric beds can result in premature discharge policies, where patients are discharged home before full recovery, leading to an increased risk of readmission (19). Our analysis showed that shorter lengths of stay were associated with an increased risk of readmission at specialized hospitals, but up to a certain point only, with lengths of stay above 90 days not decreasing the risk of readmission. Interestingly longer lengths of stay at district/acute hospitals were associated with an increased risk of readmission, possibly suggesting that these patients might benefit from specialized care. At the same time direct admissions to specialized hospitals had higher aORs for readmission, highlighting the importance of patients being managed at the appropriate level of care.

In 2022, 40% of admitted psychiatric patients had not been seen at PHC facility in the preceding year (a small proportion of these patients might still be in care, if they are being followed up by private facilities, hospital out-patient departments, or community health workers). The assumption that if these patients were regularly being followed up at the PHC level a hospital admission could be averted might not always be true, but by not following these patients up, we are potentially missing an opportunity to intervene. The relatively stable median days between PHC visit and psychiatric hospital admission of 35 to 42 days, despite the increasing proportion of individuals visiting a PHC in the year prior to admission, reiterates this. Presumably patients receive their medication for 28 days, but do not return after, resulting in an exacerbation of symptoms and admission soon after.

And while out-patient services were impacted by COVID-19, particularly in early 2020, a finding which is supported by data from the South African private sector (20), the overall trend was of increasing, albeit slowly, PHC use prior to admission. However, it is important to note that digitized PHC headcount data became mandatory for provincial PHC facilities in April 2020, and so at least some of the increase we are seeing is related to better electronic data capture rather than an actual change in PHC usage.

With regards to linkage post-discharge, male patients and those with substance related diagnoses were less likely be linked after discharge, at both hospital levels. This is in line with data from a study in patients with schizophrenia in Cape Town that found that substance use disorders were most strongly associated with non-adherence (21). Ongoing psychotic symptoms, impaired insight and involvement with the criminal justice system have been found to be barriers to engagement with ambulatory services (22). Those with longer hospital admissions were also more likely to be successfully linked to care, possibly reflecting that those individuals have more severe disease and/or that during the longer hospital stay more time was put into preparing post-discharge care. Later admission years were associated with more successful linkage to care, although improving electronic PHC headcount data might be biasing this as well.

Successful linkage to care after discharge was one of the most protective factors against readmission within 30 and 90 days, at both hospital levels. All the other covariates in these analyses are relatively fixed, with the health system having limited control over them, the exception being length of hospital stay - although with limited bed capacity this ability is also constrained. PHC visits in the year prior to admission were associated with an increased risk of readmission, possibly indicating the chronicity of the underlying disease more than the subsequent health seeking behaviour. Linkage to ambulatory psychiatric services remains a challenge globally (23). And in the Western Cape, this challenge is not limited to psychiatry services only. Patients diagnosed with tuberculosis in hospital, for instance, are more likely to be lost to follow up and have higher mortality than those diagnosed at clinics (24). Effective clinical bridging strategies proposed have included communicating discharge plans to outpatient clinicians, family involvement during the hospital stay and starting outpatient programs before discharge (23).

## Limitations

By using routinely collected electronic data, a limitation of this study is firstly that we were restricted to admitted patients only, and secondly, that we were dependent on diagnostic coding that was not always accurate or sufficiently detailed. Also because of the challenges with diagnostic coding, we could only identify patients by the admitting specialty of psychiatry, excluding psychiatric patients in Emergency Centres and those admitted to other specialties. While this will underestimate the total number of psychiatric patients in hospital, particularly for suicidal patients, the trends are still important, and the absolute numbers of patients admitted to psychiatric wards remain crucial to those services. In addition, with the expansion of electronic data use at PHC level during the study period, it is impossible to determine how much of the increase in PHC use that is related to better data capturing. Further detailed studies assessing the mental health burden at PHCs and Emergency Centres are needed.

For the readmission analyses, data on out-of-facility deaths and migration out of the province is not currently available at the PHDC. If after hospital discharge, an individual moves back to the family home in another province and relapses there, they will not be counted as a readmission, artificially lowering the readmission rate. And if there is differential out-migration by hospital drainage area, this would contribute to the varying facility readmission rates. Private patients also access psychiatric services in the public sector, but billing information was too incomplete to be adjusted for. While the stratification of these analyses by hospital level is reasonable, it does not consider the fact that patients often move between the two levels over many years, and more comprehensive longitudinal analyses taking into account the individual patient journeys through the health system are needed.

## Conclusion

Driven by poverty and substance use, psychiatric admissions have increased markedly, particularly at the district/acute hospital level, and have been exacerbated further by the COVID-19 pandemic. While ambulatory service use before and after a psychiatric hospital admission is slowly increasing with time, more can possibly be done at that level to avert hospital admissions. Successful linkage to ambulatory services following hospital discharge is one of the most protective factors against readmission, and stronger measures need to be put in place to ensure that all individuals discharged home receive appropriate follow-up.

## Funding

Funding for this study was provided by the Western Cape Department of Health and Wellness, as well as by grants from the Swiss National Science Foundation (193381) and from the National Institutes of Health for the International Epidemiology Databases to Evaluate AIDS Southern Africa Collaboration (IeDEA-SA) (U01 AI069924).

## Data Availability

The underlying data are routinely collected patient records that have been de-identified and pseudo-anonymised. The patients have not consented to these data being part of publicly accessible repositories considering the inherent risks of re-identification.The Western Cape Department of Health and Wellness evaluates research proposals for all research in the public health sector in the province, subject to standard research ethics, government approval and data governance prescripts. This includes those that draw on routine datasets like the current study. For more information.

---

**Supplementary Figure 1: Median age in years of admitted psychiatric patients by diagnosis category and sex, in all facilities in the Western Cape 2015-2022.**

**Supplementary Figure 2: Median length of stay (LOS) in days for acute/district admissions (2a), by destination category (discharged directly home vs transferred on for specialized care), and LOS for specialized hospitals by diagnosis category (2b)**

**Supplementary Figure 3: Primary health care visits preceding an district/acute psychiatric hospital admission: proportion of admissions with a PHC visit in the 30 days prior to admission, in 31-365 days prior admission and those without a PHC visit, for 2015-2022 (3a), and proportion of admissions with a PHC visit in the 30 days prior to admission in January 2020 – December 2021 (3b).**

**Supplementary Table 1:**
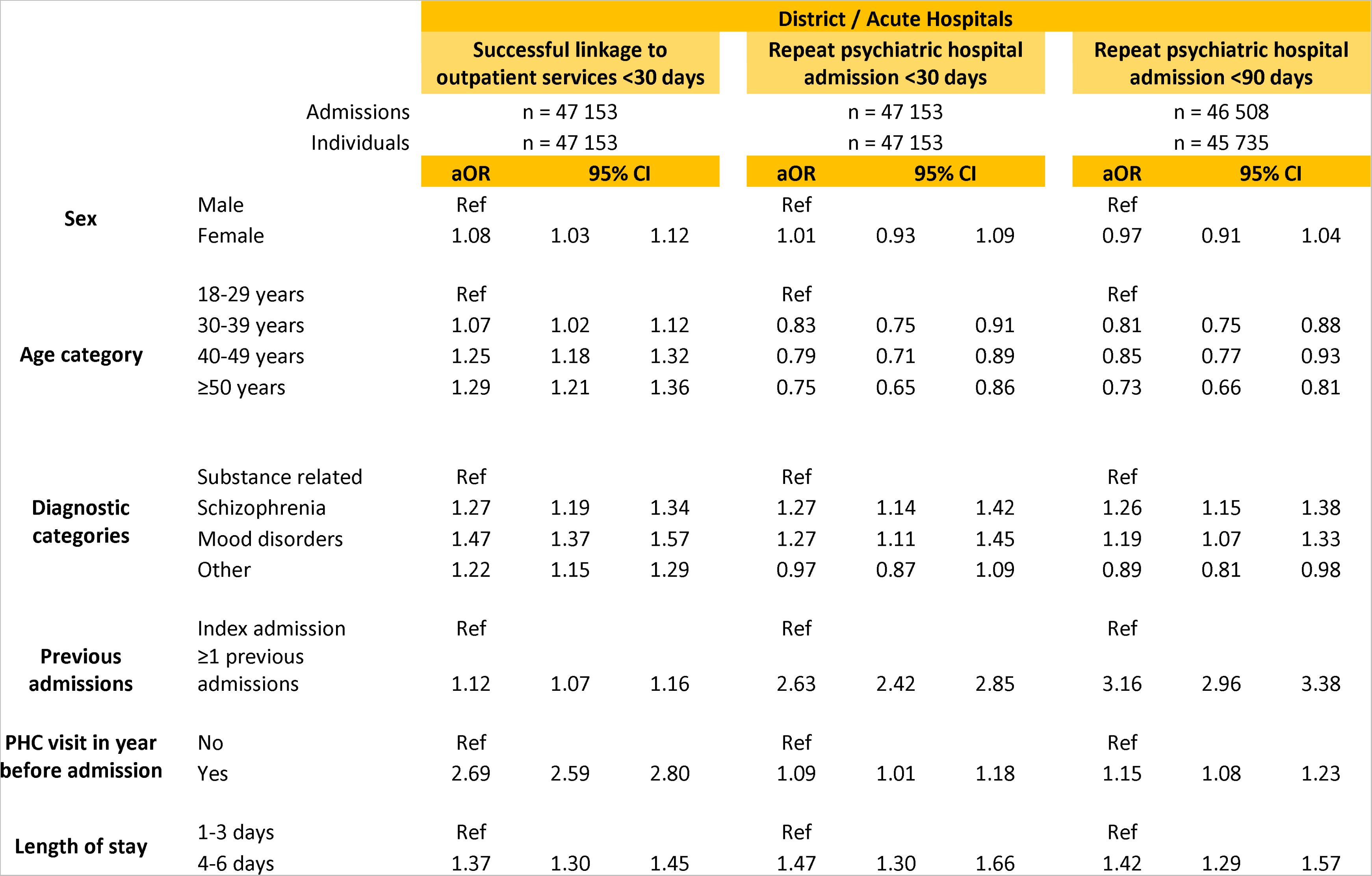
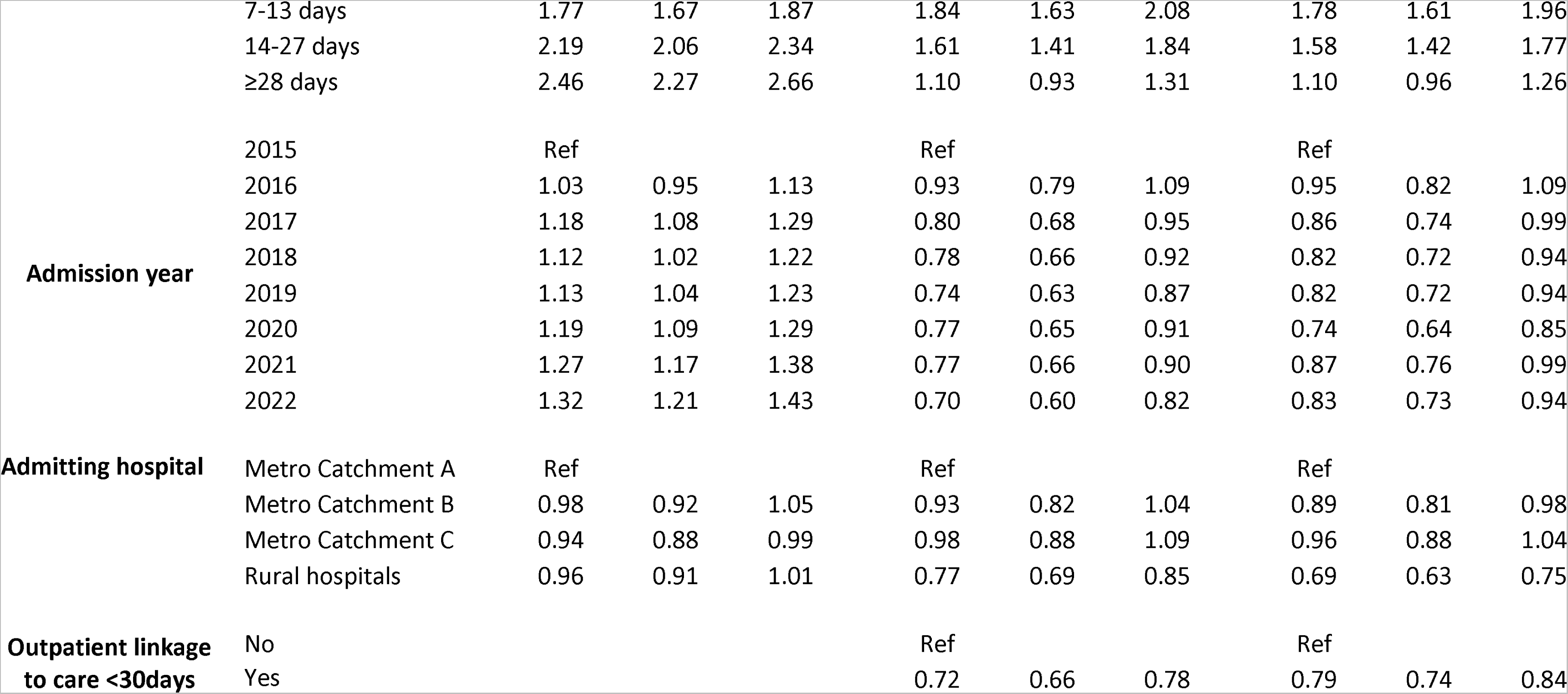
Standard logistic regression for the outcomes of successful linkage to care and psychiatric readmission within 30 and 90 days for district/acute hospitals. Only the latest admission for an individual was included in this analysis.

|  |  | District / Acute Hospitals |  |  |  |  |  |  |  |  |
| --- | --- | --- | --- | --- | --- | --- | --- | --- | --- | --- |
|  |  | Successful linkage to outpatient services <30 days |  |  | Repeat psychiatric hospital admission <30 days |  |  | Repeat psychiatric hospital admission <90 days |  |  |
|  |  | Admissions<br>n = 47 153 |  |  | n = 47 153 |  |  | n = 46 508 |  |  |
|  |  | Individuals<br>n = 47 153 |  |  | n = 47 153 |  |  | n = 45 735 |  |  |
|  |  | aOR | 95% CI |  | aOR | 95% CI |  | aOR | 95% CI |  |
| Sex | Male | Ref |  |  | Ref |  |  | Ref |  |  |
|  | Female | 1.08 | 1.03 | 1.12 | 1.01 | 0.93 | 1.09 | 0.97 | 0.91 | 1.04 |
| Age category | 18-29 years | Ref |  |  | Ref |  |  | Ref |  |  |
|  | 30-39 years | 1.07 | 1.02 | 1.12 | 0.83 | 0.75 | 0.91 | 0.81 | 0.75 | 0.88 |
|  | 40-49 years | 1.25 | 1.18 | 1.32 | 0.79 | 0.71 | 0.89 | 0.85 | 0.77 | 0.93 |
|  | ≥50 years | 1.29 | 1.21 | 1.36 | 0.75 | 0.65 | 0.86 | 0.73 | 0.66 | 0.81 |
| Diagnostic categories | Substance related | Ref |  |  | Ref |  |  | Ref |  |  |
|  | Schizophrenia | 1.27 | 1.19 | 1.34 | 1.27 | 1.14 | 1.42 | 1.26 | 1.15 | 1.38 |
|  | Mood disorders | 1.47 | 1.37 | 1.57 | 1.27 | 1.11 | 1.45 | 1.19 | 1.07 | 1.33 |
|  | Other | 1.22 | 1.15 | 1.29 | 0.97 | 0.87 | 1.09 | 0.89 | 0.81 | 0.98 |
| Previous admissions | Index admission | Ref |  |  | Ref |  |  | Ref |  |  |
|  | ≥1 previous admissions | 1.12 | 1.07 | 1.16 | 2.63 | 2.42 | 2.85 | 3.16 | 2.96 | 3.38 |
| PHC visit in year before admission | No | Ref |  |  | Ref |  |  | Ref |  |  |
|  | Yes | 2.69 | 2.59 | 2.80 | 1.09 | 1.01 | 1.18 | 1.15 | 1.08 | 1.23 |
| Length of stay | 1-3 days | Ref |  |  | Ref |  |  | Ref |  |  |
|  | 4-6 days | 1.37 | 1.30 | 1.45 | 1.47 | 1.30 | 1.66 | 1.42 | 1.29 | 1.57 |

|  |  |  |  |  |  |  |  |  |  |  |  |
| --- | --- | --- | --- | --- | --- | --- | --- | --- | --- | --- | --- |
|  |  | 7-13 days | 1.77 | 1.67 | 1.87 | 1.84 | 1.63 | 2.08 | 1.78 | 1.61 | 1.96 |
|  |  | 14-27 days | 2.19 | 2.06 | 2.34 | 1.61 | 1.41 | 1.84 | 1.58 | 1.42 | 1.77 |
|  |  | ≥28 days | 2.46 | 2.27 | 2.66 | 1.10 | 0.93 | 1.31 | 1.10 | 0.96 | 1.26 |
|  | Admission year | 2015 | Ref |  |  | Ref |  |  | Ref |  |  |
|  |  | 2016 | 1.03 | 0.95 | 1.13 | 0.93 | 0.79 | 1.09 | 0.95 | 0.82 | 1.09 |
|  |  | 2017 | 1.18 | 1.08 | 1.29 | 0.80 | 0.68 | 0.95 | 0.86 | 0.74 | 0.99 |
|  |  | 2018 | 1.12 | 1.02 | 1.22 | 0.78 | 0.66 | 0.92 | 0.82 | 0.72 | 0.94 |
|  |  | 2019 | 1.13 | 1.04 | 1.23 | 0.74 | 0.63 | 0.87 | 0.82 | 0.72 | 0.94 |
|  |  | 2020 | 1.19 | 1.09 | 1.29 | 0.77 | 0.65 | 0.91 | 0.74 | 0.64 | 0.85 |
|  |  | 2021 | 1.27 | 1.17 | 1.38 | 0.77 | 0.66 | 0.90 | 0.87 | 0.76 | 0.99 |
|  |  | 2022 | 1.32 | 1.21 | 1.43 | 0.70 | 0.60 | 0.82 | 0.83 | 0.73 | 0.94 |
|  | Admitting hospital | Metro Catchment A | Ref |  |  | Ref |  |  | Ref |  |  |
|  |  | Metro Catchment B | 0.98 | 0.92 | 1.05 | 0.93 | 0.82 | 1.04 | 0.89 | 0.81 | 0.98 |
|  |  | Metro Catchment C | 0.94 | 0.88 | 0.99 | 0.98 | 0.88 | 1.09 | 0.96 | 0.88 | 1.04 |
|  |  | Rural hospitals | 0.96 | 0.91 | 1.01 | 0.77 | 0.69 | 0.85 | 0.69 | 0.63 | 0.75 |
|  | Outpatient linkage to care <30days | No |  |  |  | Ref |  |  | Ref |  |  |
|  |  | Yes |  |  |  | 0.72 | 0.66 | 0.78 | 0.79 | 0.74 | 0.84 |

**Supplementary Table 2:**
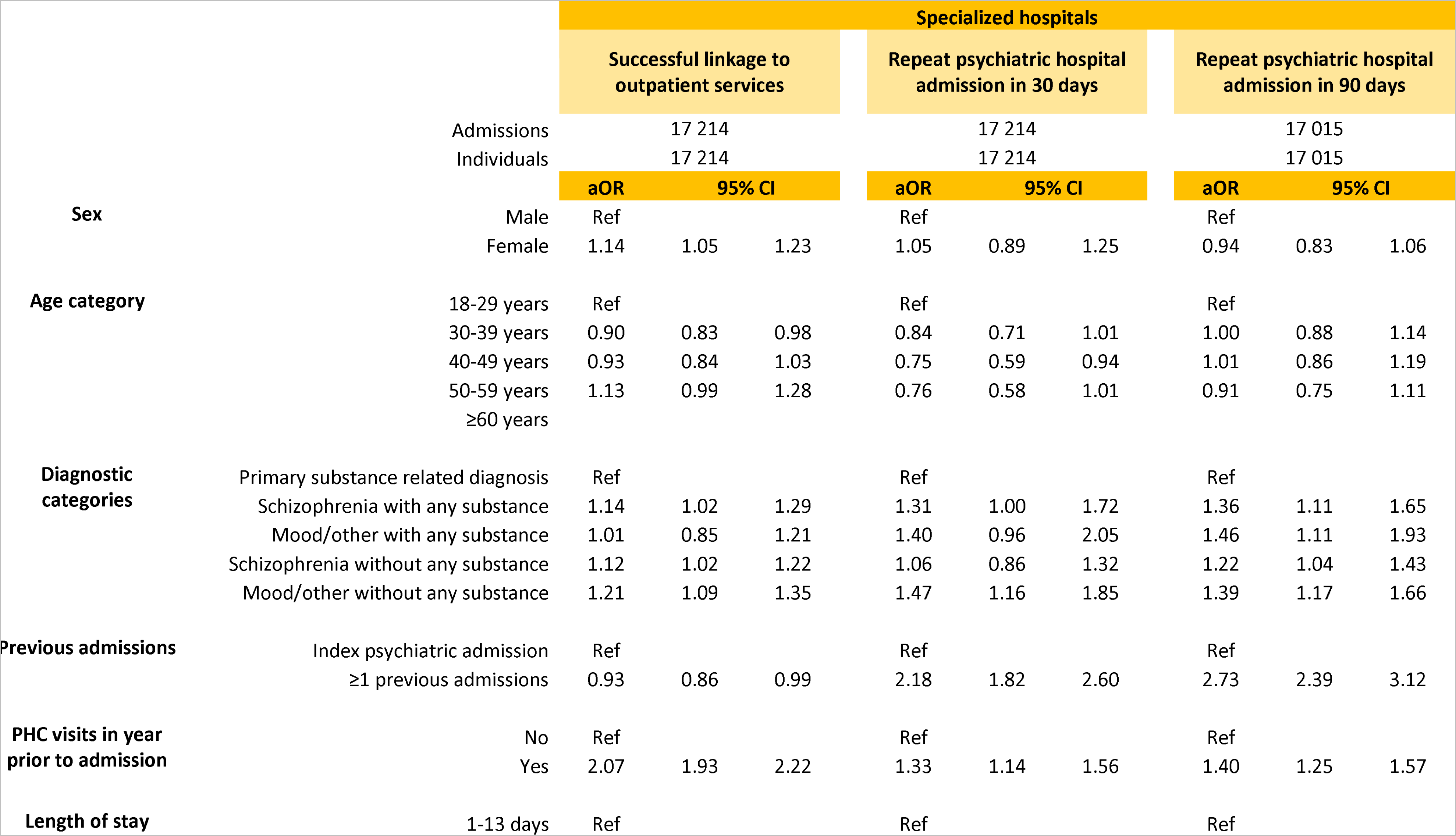
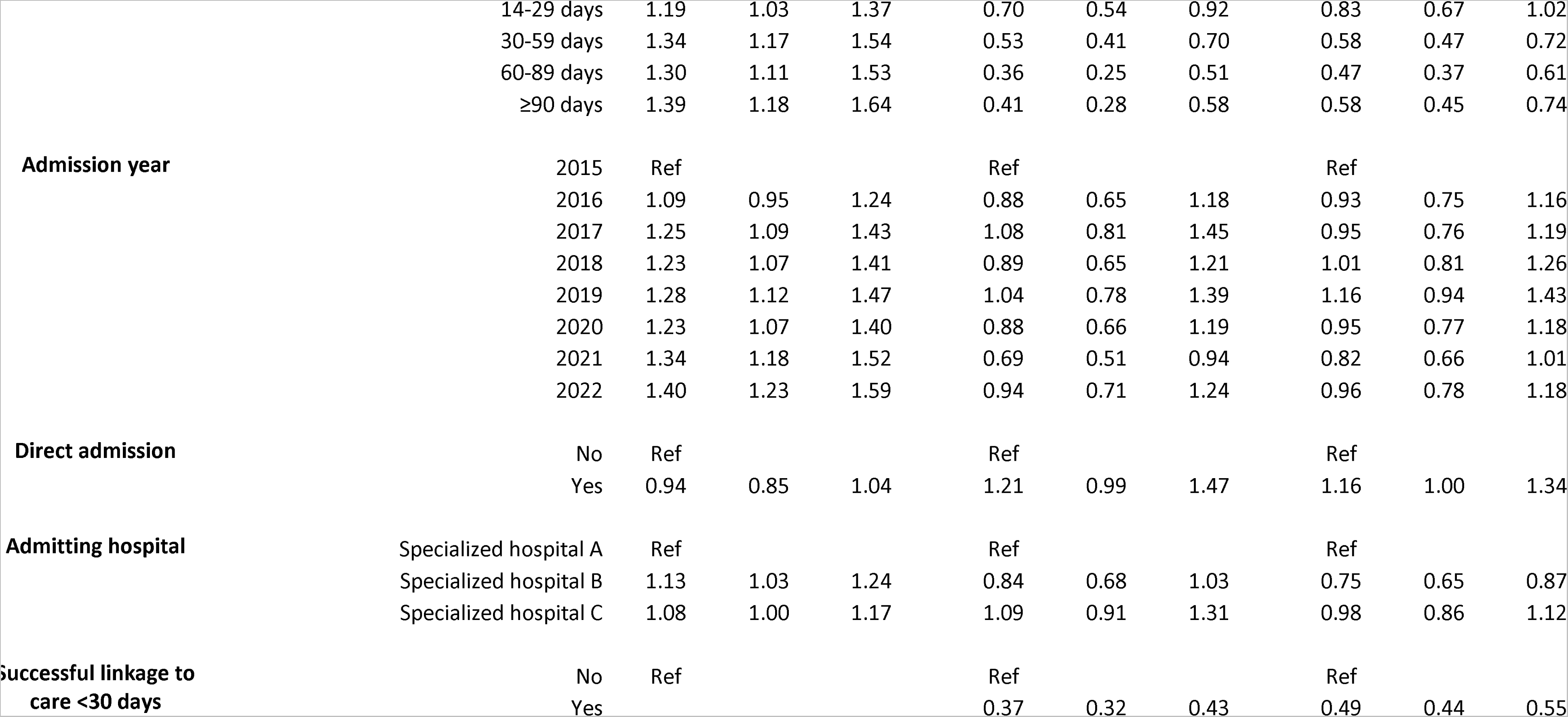
Standard logistic regression for the outcomes of successful linkage to care and psychiatric readmission within 30 and 90 days for specialized hospitals. Only the latest admission for an individual was included in this analysis.

## Notes

### Competing Interest Statement

The authors have declared no competing interest.

### Author Declarations

This study was approved by the University of Cape Town Health Research Ethics Committee (HREC 058 /2023).

### Summary of Updates

This version of the manuscript has been updated to reflect the correct funding information and edited down for word count.

