## Supplementary figures and images for "Psychiatric hospital admissions and linkages to ambulatory services in the Western Cape Province of South Africa (2015-2022): trends, risk factors and possible opportunities for intervention"

### Supplementary Figure 1

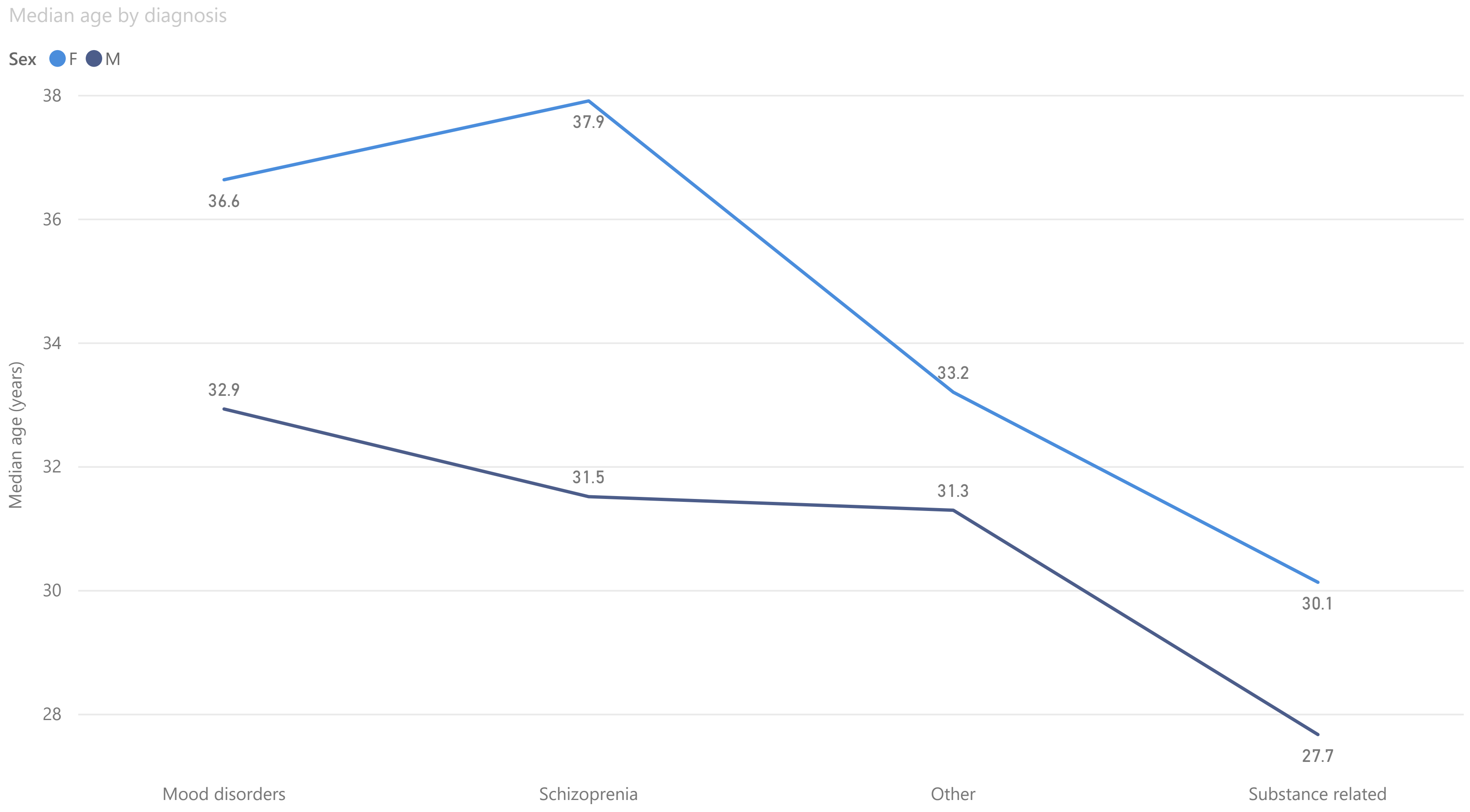
