## Supplementary Figure 2 for "Psychiatric hospital admissions and linkages to ambulatory services in the Western Cape Province of South Africa (2015-2022): trends, risk factors and possible opportunities for intervention"

Median LOS in district/acute hospitals

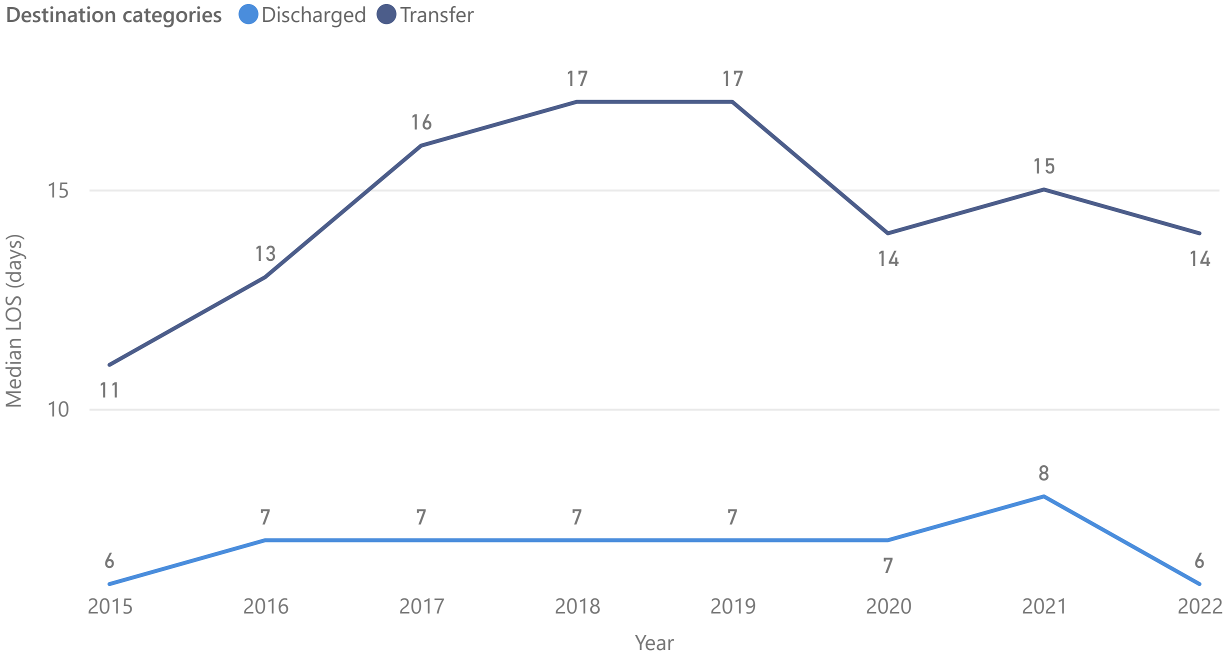


Median LOS in specialized hospitals by diagnosis category

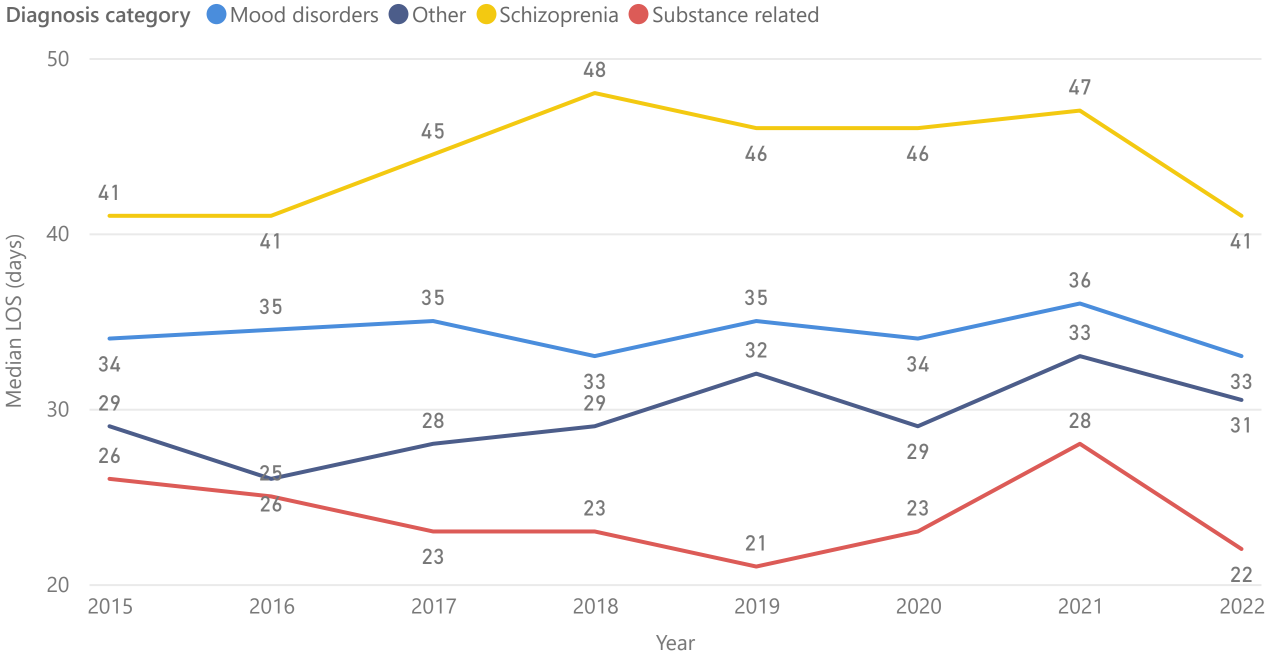
