## Supplementary Figure 3 for "Psychiatric hospital admissions and linkages to ambulatory services in the Western Cape Province of South Africa (2015-2022): trends, risk factors and possible opportunities for intervention"

PHC visits prior to admission at district/acute hospitals

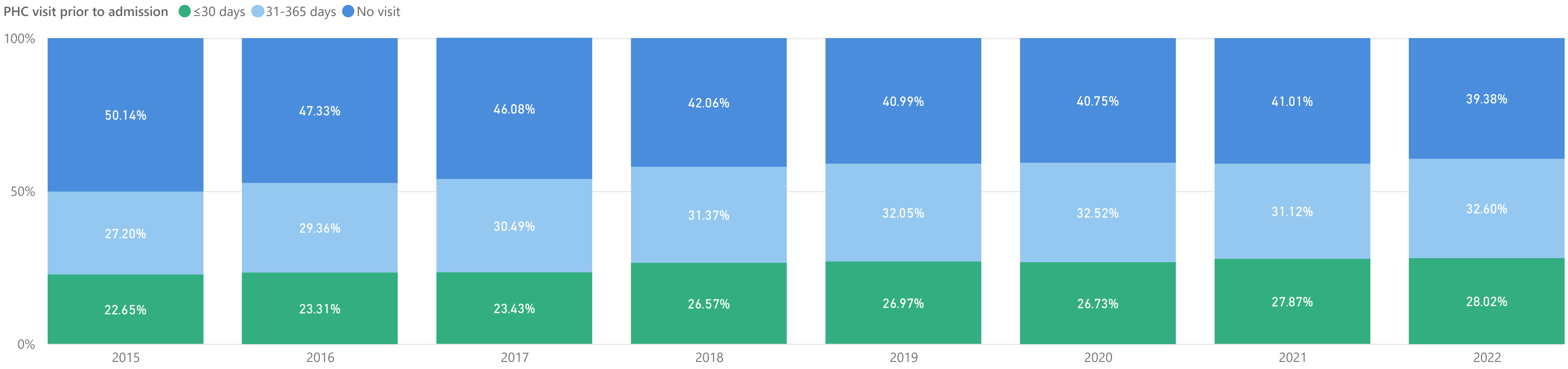


Proportion of admissions with PHC visit in 30 days prior to admission (2020-2021)

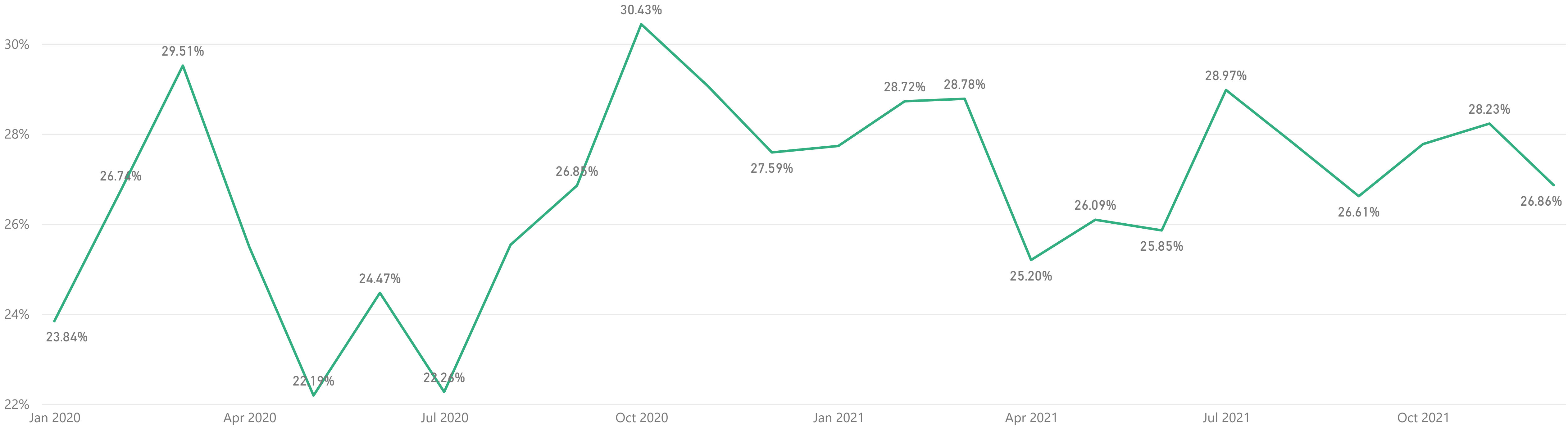
